# People with non-hepatic steatosis combined with metabolic dysfunction as defined by atherosclerotic plaques are at higher risk of early liver fibrosis

**DOI:** 10.1101/2025.04.13.25325759

**Authors:** Yujin Li, Zheyu Li, Shaochen Su, Xiaoqin Gao, Dan Zhou, Min Li, Longzhen Shi, Xiaojin Zhou, Yu Chen, Junfeng Li

**Affiliations:** Fuxing Hospital, School of Basic Medical Sciences, Capital Medical University, Beijing, China; First Clinical Medical College of Lanzhou University, Lanzhou, Gansu, China; Physical Examination Center, First Hospital of Lanzhou University, Lanzhou, Gansu, China; Department of Hepatology, First Hospital of Lanzhou University, Lanzhou, Gansu, China; Intractable Hepatic Diseases and Artificial Liver Treatment, Beijing Youan Hospital, Capital Medical University, Beijing, China; Institute of Infectious Diseases, First Hospital of Lanzhou University, Lanzhou, Gansu, China

**Author notes:** Corresponding Author: E-mail address (YC), Email address (JL). Fundings: National Natural Science Foundation of China (82360132), Major Project of Scientific and Technological Innovation in Gansu Health Industry (GSWSZD2024 - 11), Joint Scientific Research Foundation Project of Gansu Province (23JRRA1489, 24JRRA911); Key Talent Project of Gansu Province (No. [2024]4); Open Project of Key Laboratory of Extreme Environment Microbiology in Gansu Province (EEMRE202401).

**Keywords:** Metabolic dysfunction, hepatic steatosis, hepatic fibrosis

## Abstract

**Objective:** This study examines the association between hepatic steatosis (HS) and liver fibrosis risk in individuals with similar metabolic disorders but differing metabolic dysfunction indicators.

**Methods:** A cross-sectional study was conducted at the First Hospital of Lanzhou University from January to November 2024, involving individuals undergoing physical examinations. Laboratory tests, vascular and abdominal ultrasound examinations, and clinical data were collected to assess metabolic characteristics. Logistic regression models were used to explore the relationship between metabolic dysfunction, HS, and liver fibrosis degree.

**Results:** A total of 4,006 patients were included, with 1,164 (29.06%) females and 2,157 (53.84%) having HS. Univariate logistic regression identified HS as a risk factor for early liver fibrosis (odds ratio [OR]: 1.67, 95% CI: 1.47–1.89, P < 0.001). However, after adjusting for metabolic dysfunction indicators, the correlation between HS and liver fibrosis risk weakened or reversed. Multivariate analysis, accounting for confounders, found HS to be a protective factor (OR: 0.57, 95% CI: 0.45–0.71, P < 0.001), while body roundness index (OR: 1.62, 95% CI: 1.32– 2.00, P < 0.001) and hyperglycemia (OR: 13.28, 95% CI: 10.76–16.39, P < 0.001) were identified as risk factors. Propensity score matching revealed 880 matched pairs of patients, showing that among those with atherosclerotic plaques (OR: 0.58, 95% CI: 0.42–0.79, P < 0.001) or hyperglycemia (OR: 0.63, 95% CI: 0.43–0.93, P = 0.02), HS was associated with a reduced risk of early liver fibrosis. The non-HS group had the strongest ability to detect early liver fibrosis (AUC: 0.57, 95% CI: 0.52–0.61, P = 0.003).

**Conclusion:** Metabolic dysfunction is positively correlated with the risk of liver fibrosis. However, in individuals with metabolic dysfunction, particularly those with atherosclerotic plaque formation, the non-HS population is at a higher risk of early liver fibrosis.

**Financial support:** National Natural Science Foundation of China (82360132), Major Science and Technology Innovation Project of Gansu Provincial Health Industry (GSWSZD2024-11), Joint Scientific Research Fund of Gansu Province (23JRRA1489, 24JRRA911), Key Talent Project of Gansu Province (GanZuTongZi [2024] No.4), Open Project of Gansu Provincial Key Laboratory of Extreme Environment Microbiology (EEMRE202401), Key Research and Development Program of Gansu Province (22YF7FA085), Key Project of Traditional Chinese Medicine in Gansu Province (GZKZ-2022-7), Lanzhou Science and Technology Bureau Project (2023-2-76), and Medical Education Development Projects of Lanzhou University (lzuyxcx-2022-131; lzuyxcx-2022-213; lzuyxcx-2022-147).

## Introduction

Research has demonstrated that the prevalence of hepatic steatosis (HS) in China has reached 44.39% [1]. Non-alcoholic fatty liver disease (NAFLD), primarily characterized by HS, has gradually become the leading cause of chronic liver disease globally, with its prevalence increasing annually [2]. China has the fastest growth rate and the youngest age of onset for NAFLD [2], suggesting that as the population ages, the disease burden associated with NAFLD increases significantly. Steatotic liver disease (SLD) was introduced in 2023 as a new nomenclature [3] to more accurately describe liver diseases predominantly characterized by HS, replacing NAFLD. SLD encompasses metabolic dysfunction-associated steatotic liver disease (MASLD), MASLD with increased alcohol intake, alcohol-associated liver disease, or etiologically specific/cryptogenic SLD [3].

Conventionally, HS was not considered a strong predictor of liver fibrosis progression in NAFLD [4]. However, recent studies demonstrated that HS had a significant predictive value for liver fibrosis progression in SLD. Different studies reported inconsistent findings [5-8]. In the early stages of SLD, increased HS is associated with liver fibrosis progression [5]. However, when significant liver fibrosis occurs, HS is no longer prominent in the SLD population [9]. This may explain the inconsistent findings regarding the prognostic value of liver steatosis in various studies. However, this hypothesis does not emphasize the role of metabolic dysfunction in liver fibrosis progression in SLD. First, metabolic dysfunction is closely associated with HS [1], a local manifestation of metabolic syndrome. Second, metabolic dysfunction is associated with liver fibrosis progression in SLD [1, 10], and its predictive value for adverse outcomes may even be higher than that of HS [11]. Furthermore, multiple criteria exist for metabolic dysfunction, and different metabolic abnormality indicators may have varying risks for liver fibrosis in SLD [12]. Therefore, neglecting the association between metabolic dysfunction and HS, its potential role in liver fibrosis progression, or disregarding its heterogeneity in studies on the poor prognosis of SLD associated with HS may result in inaccurate research conclusions.

Some studies only include people with HS. However, due to the current unclear association between HS and the risk of liver fibrosis, excluding non-HS populations may result in inconsistencies in research conclusions. Consequently, it is essential to investigate further the association between HS and the risk of liver fibrosis in HS and non-HS populations, including individuals undergoing routine physical examinations. Simultaneously, attention must be directed toward the impact of metabolic dysfunction and its heterogeneity on research results.

This study aimed to investigate the association between HS and the risk of liver fibrosis in a large population undergoing medical examination, considering various metabolic dysfunction indicators under similar degrees of metabolic disorders.

## Materials & Methods

### 1. Study Subjects and Observation Indicators

This cross-sectional study screened and collected clinical data from all individuals who participated in physical examinations at the First Hospital of Lanzhou University’s medical examination center from January 2024 to November 2024. This project was approved by the First Hospital of Lanzhou University Ethics Committee (Approval No. LDYYLL2025-103). Exclusion criteria included the following: (1) Patients aged < 18 years old; (2) patients with incomplete or missing general, laboratory, or imaging data. The demographic data included gender, age, height, weight, blood pressure, and waist circumference. The laboratory data included fasting blood glucose (GLU), triglycerides (TG), high-density lipoprotein (HDL), low-density lipoprotein (LDL), platelets (PLT), alanine aminotransferase (ALT), aspartate aminotransferase (AST), albumin (ALB), and direct bilirubin (DBIL). The imaging data comprised abdominal ultrasound and carotid ultrasound. This study is a retrospective analysis of case data and has been granted an exemption from informed consent requirements. It was approved by the First Hospital of Lanzhou University Ethics Committee (No. LDYYLL2025-103). The author did not have access to information that could identify individual participants during or after data collection.

Due to the absence of concurrent blood pressure and waist circumference data in the population, we created a waist circumference group comprising 4,006 individuals without blood pressure data and a blood pressure group comprising 3,553 individuals without waist circumference data based on the abovementioned screening. We performed correlation analyses for the groups, and the results were similar. The analysis results for the waist circumference group are presented in the main text. The results of the correlation analysis for the blood pressure group are included in the Supplementary materials.

### 2. Diagnosis of Hepatic Fibrosis, HS, and Evaluation of Metabolic Dysfunction Indicators

The NAFLD Fibrosis Score (NFS) [13] was used to evaluate hepatic fibrosis. NFS < –1.44 was classified as Stage 1; NFS ≥ 0.672 was classified as Stage 3, and the remaining values were classified as Stage 2. The risk of early hepatic fibrosis risk was established as NFS ≥ –1.44. HS and splenomegaly were diagnosed through abdominal ultrasound, and atherosclerotic plaques were diagnosed using neck vascular ultrasound.

The MASLD criteria [3] defines obesity as body mass index ≥ 23 kg/m2; hyperglycemia as GLU ≥ 5.6 mmol/L; hypertension as blood pressure ≥ 130/85 mmHg or the administration of antihypertensive medication; high TG as TG ≥ 1.7 mmol/L or the administration of lipid-lowering medication; and low HDL as HDL ≤ 1.0 mmol/L/1.3 mmol/L (male/female) or the administration of lipid-lowering medication. Central obesity is defined as ≥ 94 cm/80 cm (male/female) based on the metabolic syndrome criteria [14].

The scoring models for metabolic dysfunction indicators included: METS-IR [15], TyG [16], TyG-BMI [17], TyG-WC [18], TyG-WtHR [19], HSI [20], VAI [21], LAP [22], ZJU [23], and FSI [24]. The defined metabolic dysfunctions were: METS-IR ≥ 35.223, TyG ≥ 8.680, TyG-BMI ≥ 211.515, TyG-WC ≥ 714.871, TyG-WtHR ≥ 4.198, HSI ≥ 33.032, VAI ≥ 1.426, LAP ≥ 28.720, ZJU ≥ 34.549, and FSI ≥ –2.408 [25].

### 3. Statistical Methods

Statistical Package for the Social Sciences software (version 26.0) was used for data analysis. Skewed distribution measurement data are expressed as M(P25-P75), and the Kruskal– Wallis test was used to compare groups. The chi-square test was used to compare count data between groups. A binary logistic regression model was utilized to analyze the correlation between relevant indicators and the risk of early hepatic fibrosis. To further eliminate the effect of metabolic dysfunction on the association between HS and hepatic fibrosis, propensity score matching (PSM) was performed at a 1:1 ratio for two groups with and without HS. Matching covariates for the waist circumference group included gender, age, body roundness index (BRI), BMI, waist circumference, obesity, central obesity, GLU, hyperglycemia, triglyceride, high TG, HDL, low HDL, and LDL. Matching covariates for the blood pressure group included gender, age, BMI, obesity, systolic blood pressure, diastolic blood pressure, hypertension, fasting blood glucose, hyperglycemia, triglyceride, high TG, HDL, low HDL, and LDL, with a caliper value of 0.02. Receiver operating characteristic (ROC) curves were generated, and the area under the ROC curve (AUC) was compared to analyze the ability of non-HS individuals to identify early hepatic fibrosis under different metabolic dysfunction indicators. A P < 0.05 was considered statistically significant.

**Fig 1.**
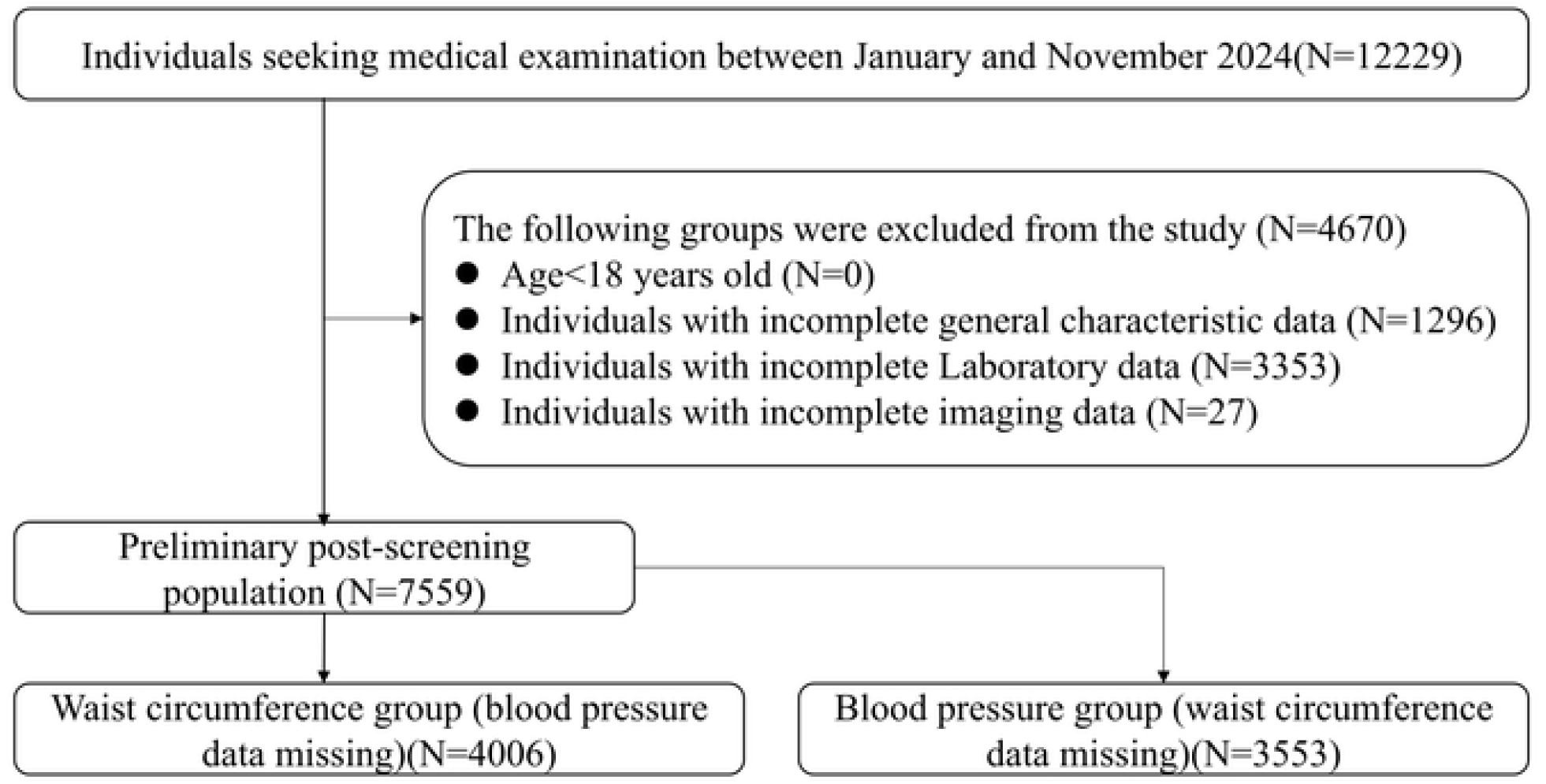
Flowchart for Screening of the Medical Examination Population.

## Results

### 1. Characteristics of the Overall Study Population

This study included 4,006 individuals, of which 1,164 (29.06%) were females. The median age of the participants was 53 (44–60.00 years), and 2,157 individuals (53.84%) were diagnosed with HS (Table 1). The HS population exhibited a higher age, NFS scores, increased obesity, central obesity, hyperglycemia, high TG, low HDL, plaque formation, and splenomegaly, and a lower proportion of females (P < 0.05) (Table 1).

**Table 1.** Characteristics of the overall population.

| Variables | Total(n=4006) | Non-HS(n=1849) | HS(n=2157) | Statistic | P |
| --- | --- | --- | --- | --- | --- |
| Age (years) | 53.00(44.00,60.00) | 51.00(42.00,60.00) | 54.00(46.00,61.00) | Z=-5.67 | <.001 |
| Female | 1164(29.06) | 773(41.81) | 391(18.13) | $\chi^2=270.81$ | <.001 |
| BMI (kg/m<sup>2</sup>) | 24.52(22.41,26.56) | 22.66(20.89,24.44) | 25.91(24.39,27.74) | Z=-35.66 | <.001 |
| Waist circumference (cm) | 87.50(80.00,93.00) | 81.00(74.00,87.00) | 91.00(87.00,96.00) | Z=-34.39 | <.001 |
| BRI | 3.64(2.98,4.27) | 3.08(2.50,3.64) | 4.05(3.57,4.64) | Z=-34.26 | <.001 |
| GLU (mmol/L) | 5.31(4.99,5.80) | 5.16(4.88,5.53) | 5.45(5.11,6.05) | Z=-16.53 | <.001 |
| TG (mmol/L) | 1.40(1.02,2.03) | 1.13(0.86,1.51) | 1.73(1.26,2.41) | Z=-26.10 | <.001 |
| HDL (mmol/L) | 1.17(1.02,1.38) | 1.30(1.11,1.50) | 1.10(0.96,1.24) | Z=-22.92 | <.001 |
| LDL (mmol/L) | 2.85(2.36,3.35) | 2.72(2.27,3.22) | 2.97(2.47,3.44) | Z=-9.31 | <.001 |
| ALT (U/L) | 23.00(17.00,33.00) | 19.00(14.00,26.00) | 27.00(20.00,38.00) | Z=-23.72 | <.001 |
| AST (U/L) | 22.00(19.00,27.00) | 22.00(18.00,25.00) | 23.00(20.00,28.00) | Z=-10.19 | <.001 |
| ALB (g/L) | 45.30(43.70,46.80) | 45.00(43.50,46.50) | 45.50(44.00,47.00) | Z=-7.69 | <.001 |
| DBIL (μmol/L) | 4.79(3.73,6.23) | 4.73(3.73,6.17) | 4.83(3.73,6.26) | Z=-0.93 | 0.355 |
| Obesity | 2756(68.80) | 834(45.11) | 1922(89.11) | $\chi^2=897.86$ | <.001 |
| Central obesity | 1419(35.42) | 319(17.25) | 1100(51.00) | $\chi^2=495.58$ | <.001 |
| Hyperglycemia | 1323(33.03) | 408(22.07) | 915(42.42) | $\chi^2=186.47$ | <.001 |
| High TG | 1457(36.37) | 338(18.28) | 1119(51.88) | $\chi^2=485.60$ | <.001 |
| Low HDL | 1319(32.93) | 439(23.74) | 880(40.80) | $\chi^2=131.12$ | <.001 |
| NFS | -1.79(-2.66,-0.84) | -1.99(-2.80,-1.09) | -1.59(-2.50,-0.69) | Z=-8.13 | <.001 |
| NFS grades | | | | $\chi^2=70.27$ | <.001 |
| 0 | 2378(59.36) | 1219(65.93) | 1159(53.73) |  |  |
| 1 | 1506(37.59) | 567(30.67) | 939(43.53) |  |  |
| 2 | 122(3.05) | 63(3.41) | 59(2.74) |  |  |
| Atherosclerotic Plaques | 1507(37.62) | 584(31.58) | 923(42.79) | $\chi^2=53.28$ | <.001 |
| Splenomegaly | 87(2.17) | 26(1.41) | 61(2.83) | $\chi^2=9.47$ | 0.002 |

### 2. Factors Associated with the Risk of Early Hepatic Fibrosis

A univariate logistic regression analysis with early hepatic fibrosis as the outcome variable revealed that HS was a risk factor for early hepatic fibrosis (odds ratio [OR]: 1.67, 95% confidence interval [CI]: 1.47–1.89, P < 0.001). When adjusting for metabolic dysfunction indicators individually, the positive correlation between HS and the risk of early hepatic fibrosis attenuated or even reversed (Table 2). Following the integration of all significant factors from the univariate analysis, multivariate logistic regression revealed that HS was a protective factor for early hepatic fibrosis (OR: 0.57, 95% CI: 0.45–0.71, P < 0.001). Conversely, age (OR: 1.17, 95% CI: 1.16–1.19, P < 0.001), BRI (OR: 1.62, 95% CI: 1.32–2.00, P < 0.001), hyperglycemia (OR: 13.28, 95% CI: 10.76–16.39, P < 0.001), and splenomegaly (OR: 3.62, 95% CI: 1.94–6.77, P < 0.001) remained as risk factors for early hepatic fibrosis (Table 3).

**Table 2.** Association between HS and early liver fibrosis in the total population.

| Model | OR(95%CI) | P |
| --- | --- | --- |
| 1 | 1.67(1.47~1.89) | <.001 |
| 2 | 1.34(1.13~1.60) | <.001 |
| 3 | 1.07(0.89~1.29) | 0.476 |
| 4 | 0.88(0.72~1.07) | 0.213 |
| 5 | 0.60(0.48~0.76) | <.001 |

Model 1: Exclusion; Model 2: Inclusion of sex, age, high TG, low HDL; Model 3: Inclusion of sex, age, high TG, low HDL, obesity; Model 4: Inclusion of sex, age, high TG, low HDL, obesity, BRI, central obesity; Model 5: Inclusion of sex, age, high TG, low HDL, obesity, BRI, central obesity, and hyperglycemia.

**Table 3.** Factors associated with early liver fibrosis in the total population.

| Variables | Univariable analysis |  | Multivariable analysis |  |
| --- | --- | --- | --- | --- |
|  | OR(95%CI) | P | OR(95%CI) | P |
| Age | 1.15(1.14~1.16) | <.001 | 1.17(1.16~1.19) | <.001 |
| Female | 0.40(0.34~0.47) | <.001 | 0.76(0.60~0.98) | 0.033 |
| BRI | 1.71(1.59~1.83) | <.001 | 1.49(1.27~1.74) | <.001 |
| Obesity | 1.74(1.51~2.01) | <.001 | 1.30(0.99~1.70) | 0.056 |
| Central obesity | 1.58(1.38~1.80) | <.001 | 1.06(0.81~1.39) | 0.66 |
| Hyperglycemia | 11.49(9.82~13.44) | <.001 | 13.28(10.76~16.39) | <.001 |
| High TG | 1.02(0.89~1.16) | 0.795 |  |  |
| Low HDL | 0.92(0.80~1.05) | 0.217 |  |  |
| HS | 1.67(1.47~1.89) | <.001 | 0.57(0.45~0.71) | <.001 |
| Atherosclerotic Plaques | 4.76(4.15~5.46) | <.001 | 0.90(0.73~1.10) | 0.298 |
| Splenomegaly | 2.10(1.37~3.24) | <.001 | 3.62(1.94~6.77) | <.001 |

### 3. Characteristics of PSM Population and Risk Factors for Early Hepatic Fibrosis

Following the adjustment of population characteristics using PSM, 880 pairs of non-HS and HS samples with similar degrees of metabolic disorder were obtained (Table 4). The HS population exhibited a higher age and incidence of plaque formation; however, NFS and its grades were lower (P < 0.05). Logistic regression analysis with early hepatic fibrosis as the outcome indicator revealed that the association between HS and early hepatic fibrosis was statistically non-significant (OR: 0.91, 95% CI: 0.76–1.11, P = 0.356) (Table 5). However, age (OR: 1.18, 95% CI: 1.16–1.20, P < 0.001), sex (female) (OR: 0.66, 95% CI: 0.48–0.92, P = 0.013), BRI (OR: 1.62, 95% CI: 1.32–2.00, P < 0.001), hyperglycemia (OR: 9.71, 95% CI: 7.20– 13.11, P < 0.001), and high TG (OR: 0.69, 95% CI: 0.51–0.94, P = 0.017) were significantly associated with the risk of early hepatic fibrosis.

**Table 4.**
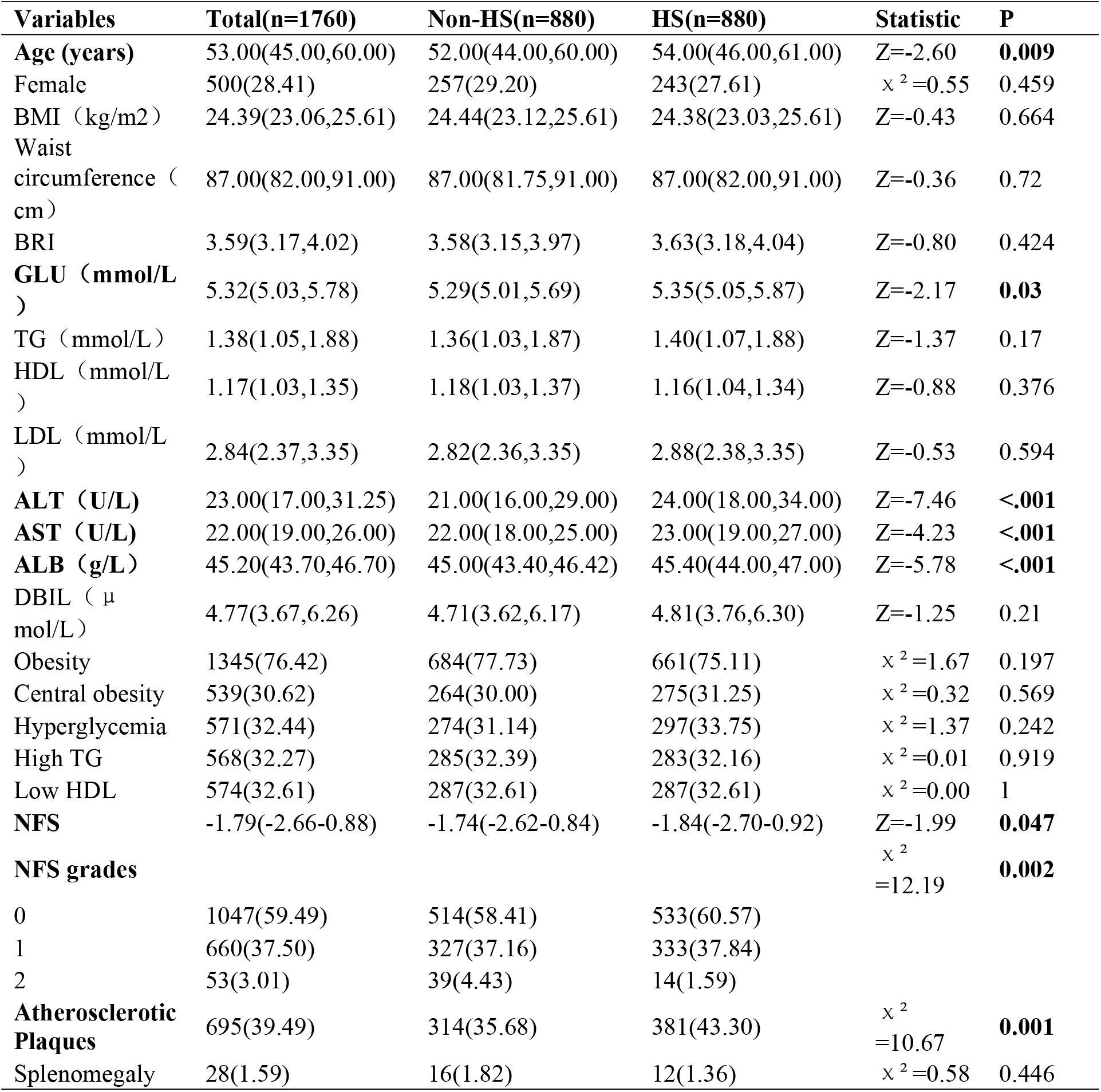
Characteristics of the PSM population.

| Variables | Total(n=1760) | Non-HS(n=880) | HS(n=880) | Statistic | P |
| --- | --- | --- | --- | --- | --- |
| **Age (years)** | 53.00(45.00,60.00) | 52.00(44.00,60.00) | 54.00(46.00,61.00) | Z=-2.60 | **0.009** |
| Female | 500(28.41) | 257(29.20) | 243(27.61) | $\chi^2=0.55$ | 0.459 |
| BMI (kg/m<sup>2</sup>) | 24.39(23.06,25.61) | 24.44(23.12,25.61) | 24.38(23.03,25.61) | Z=-0.43 | 0.664 |
| Waist circumference (cm) | 87.00(82.00,91.00) | 87.00(81.75,91.00) | 87.00(82.00,91.00) | Z=-0.36 | 0.72 |
| BRI | 3.59(3.17,4.02) | 3.58(3.15,3.97) | 3.63(3.18,4.04) | Z=-0.80 | 0.424 |
| **GLU (mmol/L)** | 5.32(5.03,5.78) | 5.29(5.01,5.69) | 5.35(5.05,5.87) | Z=-2.17 | **0.03** |
| TG (mmol/L) | 1.38(1.05,1.88) | 1.36(1.03,1.87) | 1.40(1.07,1.88) | Z=-1.37 | 0.17 |
| HDL (mmol/L) | 1.17(1.03,1.35) | 1.18(1.03,1.37) | 1.16(1.04,1.34) | Z=-0.88 | 0.376 |
| LDL (mmol/L) | 2.84(2.37,3.35) | 2.82(2.36,3.35) | 2.88(2.38,3.35) | Z=-0.53 | 0.594 |
| **ALT (U/L)** | 23.00(17.00,31.25) | 21.00(16.00,29.00) | 24.00(18.00,34.00) | Z=-7.46 | **<.001** |
| **AST (U/L)** | 22.00(19.00,26.00) | 22.00(18.00,25.00) | 23.00(19.00,27.00) | Z=-4.23 | **<.001** |
| **ALB (g/L)** | 45.20(43.70,46.70) | 45.00(43.40,46.42) | 45.40(44.00,47.00) | Z=-5.78 | **<.001** |
| DBIL (μmol/L) | 4.77(3.67,6.26) | 4.71(3.62,6.17) | 4.81(3.76,6.30) | Z=-1.25 | 0.21 |
| Obesity | 1345(76.42) | 684(77.73) | 661(75.11) | $\chi^2=1.67$ | 0.197 |
| Central obesity | 539(30.62) | 264(30.00) | 275(31.25) | $\chi^2=0.32$ | 0.569 |
| Hyperglycemia | 571(32.44) | 274(31.14) | 297(33.75) | $\chi^2=1.37$ | 0.242 |
| High TG | 568(32.27) | 285(32.39) | 283(32.16) | $\chi^2=0.01$ | 0.919 |
| Low HDL | 574(32.61) | 287(32.61) | 287(32.61) | $\chi^2=0.00$ | 1 |
| **NFS** | -1.79(-2.66-0.88) | -1.74(-2.62-0.84) | -1.84(-2.70-0.92) | Z=-1.99 | **0.047** |
| **NFS grades** | | | | $\chi^2=12.19$ | **0.002** |
| 0 | 1047(59.49) | 514(58.41) | 533(60.57) |  |  |
| 1 | 660(37.50) | 327(37.16) | 333(37.84) |  |  |
| 2 | 53(3.01) | 39(4.43) | 14(1.59) |  |  |
| **Atherosclerotic Plaques** | 695(39.49) | 314(35.68) | 381(43.30) | $\chi^2=10.67$ | **0.001** |
| Splenomegaly | 28(1.59) | 16(1.82) | 12(1.36) | $\chi^2=0.58$ | 0.446 |

**Table 5.** Factors associated with early liver fibrosis in the PSM population.

| Variables | Univariable analysis |  | Multivariable analysis |  |
| --- | --- | --- | --- | --- |
|  | OR(95%CI) | P | OR(95%CI) | P |
| Age | 1.18(1.16~1.20) | <.001 | 1.18(1.16~1.20) | <.001 |
| Female | 0.49(0.39~0.61) | <.001 | 0.66(0.48~0.92) | 0.013 |
| BRI | 1.81(1.57~2.09) | <.001 | 1.62(1.32~2.00) | <.001 |
| Obesity | 1.19(0.95~1.49) | 0.134 |  |  |
| Central obesity | 1.05(0.86~1.29) | 0.625 |  |  |
| Hyperglycemia | 9.36(7.43~11.80) | <.001 | 9.71(7.20~13.11) | <.001 |
| High TG | 0.61(0.50~0.76) | <.001 | 0.69(0.51~0.94) | 0.017 |
| Low HDL | 0.63(0.52~0.78) | <.001 | 0.98(0.72~1.33) | 0.890 |
| Atherosclerotic Plaques | 4.95(4.03~6.09) | <.001 | 0.91(0.67~1.23) | 0.528 |
| HS | 0.91(0.76~1.11) | 0.356 |  |  |
| Splenomegaly | 0.95(0.44~2.04) | 0.894 |  |  |

### 4. Correlation between HS and the Risk of Early Hepatic Fibrosis in the PSM Population with Metabolic Dysfunction

A univariate logistic regression analysis with early hepatic fibrosis as the outcome indicator revealed that in the metabolic dysfunction population defined by atherosclerotic plaques formation (OR: 0.58, 95% CI: 0.42–0.79, P < 0.001) or hyperglycemia (OR: 0.63, 95% CI: 0.43– 0.93, P = 0.02), those with HS were less prone to early hepatic fibrosis (Table 6). Additionally, in the PSM population of the blood pressure group, a lower risk of early hepatic fibrosis was observed among patients with HS in the metabolic dysfunction populations defined by atherosclerotic plaque formation (OR: 0.61, 95% CI: 0.41–0.90, P = 0.012), HSI (OR: 0.67, 95% CI: 0.49–0.91, P = 0.01), hypertension (OR: 0.69, 95% CI: 0.51–0.93, P = 0.015), ZJU (OR: 0.69, 95% CI: 0.53–0.91, P = 0.008), FSI (OR: 0.70, 95% CI: 0.55–0.90, P = 0.006), TyG-BMI (OR: 0.72, 95% CI: 0.54–0.97, P = 0.028), METS-IR (OR: 0.74, 95% CI: 0.57–0.97, P = 0.027), or obesity (OR: 0.74, 95% CI: 0.57–0.95, P = 0.020) (S1 Table).

**Table 6.**
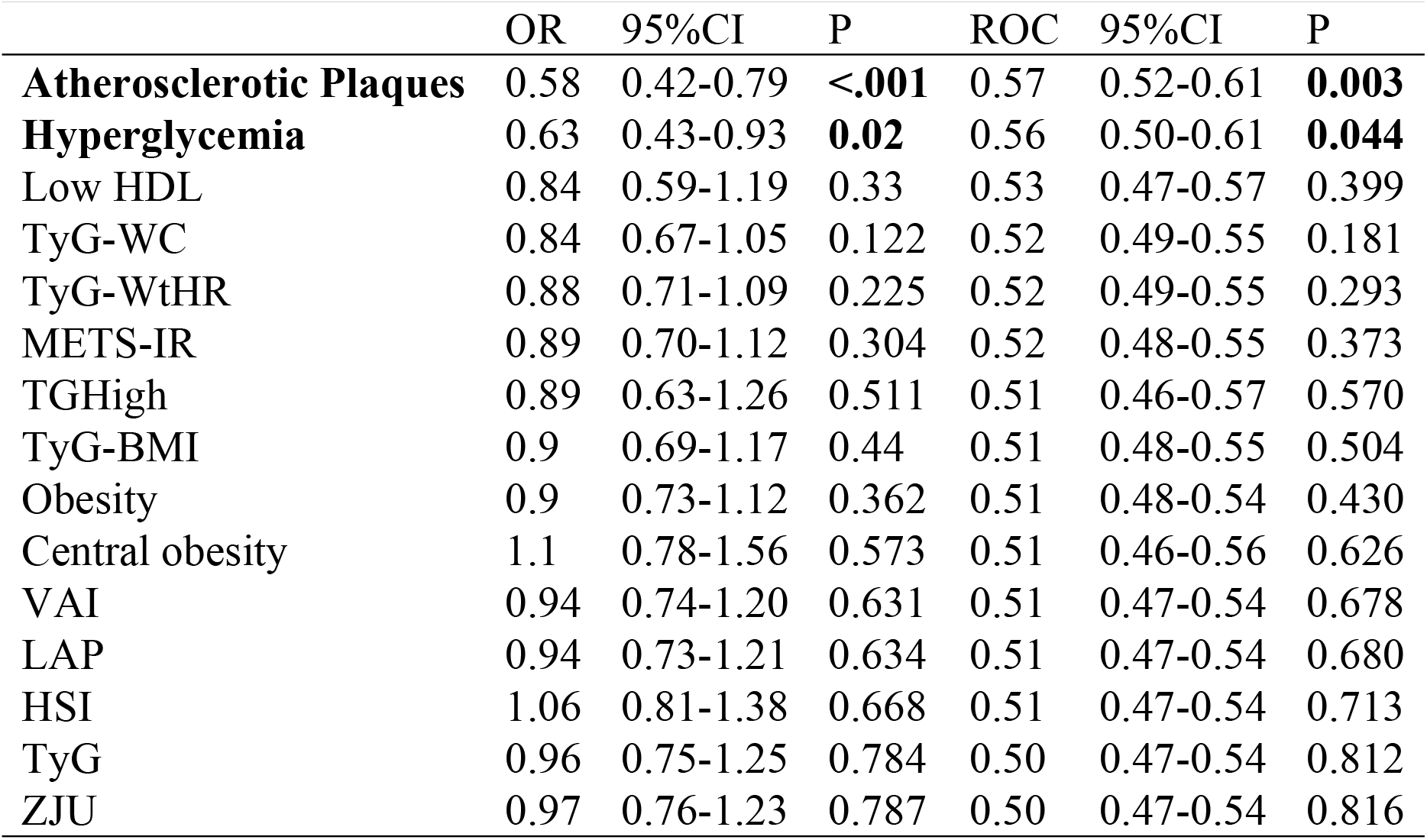
Risk of Early Hepatic Fibrosis in HS under Different Metabolic Dysfunction Indicators after PSM (Waist Circumference Group).

ROC curves were used to assess the risk of early hepatic fibrosis in HS under different metabolic function indicators. It revealed that the AUC was largest for atherosclerotic plaque formation in the waist circumference group (AUC: 0.57, 95% CI: 0.52–0.61, P = 0.003) and the blood pressure group (AUC: 0.56, 95% CI: 0.51–0.62, P = 0.03) (Table 6, S1 Table).

### 5. Other Factors Associated with Early Hepatic Fibrosis in the PSM Population with Metabolic Dysfunction

In the PSM population with metabolic dysfunction defined by atherosclerotic plaques, a multivariate logistic regression analysis revealed that age (OR: 1.19, 95% CI: 1.15–1.23, P < 0.001), BRI (OR: 1.59, 95% CI: 1.15–2.18, P = 0.005), and hyperglycemia (OR: 12.75, 95% CI: 7.90–20.60, P < 0.001) are risk factors for early hepatic fibrosis (Table 7).

**Table 7.** Factors Associated with Early Hepatic Fibrosis in the PSM Population with Atherosclerotic Plaques.

| Variables | Univariable analysis |  | Multivariable analysis |  |
| --- | --- | --- | --- | --- |
|  | OR(95%CI) | P | OR(95%CI) | P |
| **Age** | 1.16(1.13~1.19) | **<.001** | 1.19(1.15~1.23) | **<.001** |
| Female | 0.62(0.41~0.92) | **0.018** | 0.65(0.37~1.16) | 0.145 |
| **BRI** | 1.60(1.26~2.02) | **<.001** | 1.59(1.15~2.18) | **0.005** |
| Obesity | 1.08(0.75~1.55) | 0.681 |  |  |
| Central obesity | 1.21(0.86~1.71) | 0.273 |  |  |
| **Hyperglycemia** | 8.48(5.81~12.39) | **<.001** | 12.75(7.90~20.60) | **<.001** |
| High TG | 0.63(0.45~0.88) | **0.006** | 0.87(0.55~1.37) | 0.548 |
| Low HDL | 0.69(0.49~0.98) | **0.035** | 1.08(0.66~1.75) | 0.76 |
| **HS** | 0.58(0.42~0.79) | **<.001** | 0.50(0.33~0.78) | **0.002** |
| Splenomegaly | 0.48(0.15~1.59) | 0.229 |  |  |

### 6. Characteristics of the “Silent Steatotic Liver” Population with Metabolic Dysfunction but No Hepatic Steatosis

In the PSM population using atherosclerotic plaque to define metabolic dysfunction, a higher risk of hepatic fibrosis in the non-HS group was observed only in those with metabolic dysfunction (NFS grade 0 versus 1: OR: 0.65, 95% CI: 0.47–0.90, P = 0.008; 0 versus 2: OR: 0.19, 95% CI: 0.09–0.39, P < 0.001). “The population with metabolic dysfunction but no HS, termed the “silent steatotic liver” population, was more likely to occur in the elderly population (OR: 0.98, 95% CI: 0.97–0.99, P = 0.008) (Table 8).

**Table 8.** Factors Associated with Fatty Liver in the PSM Population with and without Atherosclerotic Plaque.

| Variables | Atherosclerotic Plaques |  | Non-Atherosclerotic Plaques |  |
| --- | --- | --- | --- | --- |
|  | OR(95%CI) | P | OR(95%CI) | P |
| **Age** | 0.98(0.97~0.99) | **0.008** | 1.01(1.00~1.02) | 0.079 |
| Female | 1.31(0.87~1.97) | 0.191 | 0.89(0.69~1.15) | 0.376 |
| BRI | 0.90(0.73~1.12) | 0.355 | 1.06(0.89~1.27) | 0.519 |
| Obesity | 0.78(0.55~1.12) | 0.177 | 0.92(0.69~1.22) | 0.576 |
| Central obesity | 1.06(0.76~1.47) | 0.748 | 1.08(0.84~1.40) | 0.536 |
| Hyperglycemia | 0.82(0.61~1.10) | 0.188 | 1.30(0.97~1.72) | 0.075 |
| High TG | 1.14(0.82~1.57) | 0.439 | 0.92(0.71~1.19) | 0.526 |
| Low HDL | 1.29(0.92~1.82) | 0.139 | 0.92(0.71~1.18) | 0.502 |
| **NFS grade** |  |  |  |  |
| 0 | 1.00(Reference) |  | 1.00(Reference) |  |
| 1 | 0.65(0.47~0.90) | **0.008** | 1.00(0.76~1.33) | 0.98 |
| 2 | 0.19(0.09~0.39) | **<.001** | 0.68(0.16~2.86) | 0.598 |
| Splenomegaly | 0.68(0.21~2.26) | 0.532 | 0.79(0.30~2.09) | 0.637 |

## Discussion

This is the first study based on an extensive physical examination population that investigated the association between HS and the risk of liver fibrosis under similar degrees of metabolic disorders, incorporating various indicators of metabolic dysfunction. Our results demonstrated that among patients with metabolic dysfunction, particularly those with atherosclerotic plaque formation, non-HS individuals have a higher risk of early liver fibrosis. We term this phenomenon “silent steatotic liver,” describing a condition where HS is expected but is absent.

Regarding the protective effect of HS on SLD prognosis, the significance of HS in the SLD population was lost when significant liver fibrosis occurred, suggesting that burnt-out NASH is the probable cause [26]. However, early meta-analyses displayed that in biopsy-confirmed NASH populations, lobular inflammation independently predicted liver fibrosis progression instead of metabolic dysfunction (obesity, hyperlipidemia, and diabetes)[27]. However, this conclusion was based on a multifactor regression model; thus, the effect of metabolic dysfunction and HS on liver fibrosis progression may be obscured by lobular inflammation. Notably, the overall degree of liver fibrosis demonstrated a deteriorating trend, while the degree of HS exhibited an improving trend [27], a phenomenon similar to burnt-out NASH. Similarly, recent meta-analyses revealed that among patients with NASH with significant fibrosis, those with low liver fat content are more prone to adverse outcomes [8]. However, the finding in our study that non-HS individuals have a higher risk of early liver fibrosis does not align with the concept of burnt-out NASH. This study included SLD and non-SLD populations, and the incidence of significant liver fibrosis was rarely observed in our study population.

Contradictory conclusions exist regarding the impact of HS on adverse outcomes in populations without significant liver fibrosis. In obese or overweight populations, patients with HS exhibited lower all-cause mortality and lower rates of cardiovascular adverse event rates than non-HS individuals [12], indicating that HS is a protective factor against disease progression. Similarly, in diabetic populations with mostly non-significant liver fibrosis, patients with HS have lower all-cause mortality rates than non-HS individuals [28]. Unlike the protective effect of HS on all-cause mortality, when evaluating the association between HS and the risk of liver fibrosis, studies have found that HS is associated with an increased risk of liver fibrosis. In the MASLD population with liver stiffness measurement < 2.5 kPa, severe HS predicts the progression of liver fibrosis than mild HS [5]. Although this study considered metabolic disorder factors, including obesity, hypertension, hyperlipidemia, and type 2 diabetes, it neglected the non-HS population [5]. A previous study that included non-HS individuals demonstrated that univariate analysis revealed a higher risk of hepatic fibrosis in the HS population among those who were classified as obese or overweight; however, this association became non-significant after adjusting for variables including BMI [12]. Another study [12], which included HS and non-HS populations while considering metabolic dysfunction and its heterogeneity, yielded similar results. However, the adjustment for metabolic dysfunction in the study[12] was insufficient. After excluding the lean population, MAFLD was categorized based on the presence or absence of obese and overweight individuals, resulting in metabolic healthy MAFLD and metabolic unhealthy MAFLD. In the assessment of liver fibrosis risk of metabolic unhealthy MAFLD, the control group comprised obese and overweight, rather than metabolic unhealthy non-MAFLD, leading to inconsistencies in the extent of metabolic abnormalities between HS and non-HS populations. Furthermore, in multivariate analysis, the study[12] adjusted solely for BM among metabolic dysfunction indicators. Thus, the conclusion that HS is significantly associated with an increased risk of liver fibrosis in metabolic unhealthy MAFLD may be biased. Similarly, other studies indicating that HS increases the risk of early liver fibrosis lacked adequate adjustment for metabolic dysfunction indicators [10] or excluded non-HS populations [5].

We propose that the inconsistent conclusions regarding the association between HS and liver fibrosis risk stem from insufficient consideration of metabolic dysfunction, its heterogeneity, and the neglect of non-HS populations. However, when non-HS populations are included and metabolic dysfunction and its heterogeneity are fully considered, as evidenced in our study, non-HS individuals exhibit a higher risk of liver fibrosis among those with metabolic abnormalities. A negative correlation between HS and liver fibrosis risk is observed in basic research. For instance, ablation of relevant genes to inhibit HS reduces HS in mice with methionine- and choline-deficient diet-induced MASH but exacerbates liver inflammation and fibrosis [29]. A meta-analysis has verified the protective effect of HS on liver fibrosis progression in populations with chronic HBV infection and concomitant HS [30]. HS was diagnosed using imaging or liver biopsy in the included studies[30]. Although another meta-analysis[31] published in the same year suggested that HS is a risk factor for liver fibrosis in CHB, it included a study[32] that employed serological index scoring models for HS diagnosis, which, with extensive participants, may have caused deviations in the research results. This is because although the HS diagnosed by serological scoring models is significantly correlated with imaging HS, they are not completely equivalent. Thus, utilizing serological index scoring for HS diagnosis may yield results more indicative of metabolic dysfunction rather than a correlation between HS and poor prognosis. Our study demonstrated that utilizing ZJU or FSI as metabolic dysfunction indicators facilitated the detection of early liver fibrosis risk in non-HS populations, thereby potentially corroborating the above perspective.

The highlights of our study are as follows: First, by adjusting metabolic dysfunction indicators individually, the study demonstrated the attenuation or even reversal of the effect of increased HS on liver fibrosis risk. After eliminating the interference of metabolic disorder factors, a negative correlation was found between non-HS and early liver fibrosis. Second, the study focused on the heterogeneity of metabolic dysfunction by independently analyzing different metabolic dysfunction indicators. Hyperglycemia was identified as the most significant indicator for early liver fibrosis risk, while the non-HS populations exhibit the greatest capacity to detect early liver fibrosis when metabolic dysfunction is characterized by atherosclerotic plaque formation. This study evaluates HS using imaging, preventing the interference of metabolic dysfunction assessments associated with serological index models. This study has some limitations: First, the liver fibrosis evaluation was not based on pathological biopsy but on the NFS, and the evaluation of HS was based on abdominal ultrasonography. Hence, individuals with mild HS might be missed. Second, the study population comprised individuals undergoing physical examinations, and due to limited clinical data, factors including alcohol consumption and smoking were not adjusted. Third, this cross-sectional clinical study cannot comprehensively clarify the causal relationship and mechanism between HS and liver fibrosis. Therefore, future research should utilize pathological biopsies or non-invasive techniques with enhanced correlations to liver fibrosis and design more rigorous prospective studies to further validate the association between HS and liver fibrosis and investigate its underlying mechanisms.

## Conclusions

In conclusion, metabolic dysfunction, particularly hyperglycemia, positively correlates with liver fibrosis risk in the physical examination population. In individuals with metabolic dysfunction, particularly those with atherosclerotic plaque formation, the risk of liver fibrosis is comparatively reduced in those with HS. These findings highlight the need to maintain vigilance in clinical practice, even in non-HS populations, particularly those exhibiting metabolic dysfunction. Their liver fibrosis status must be monitored, and timely intervention should be provided. A question to consider pertains to the reasons why HS is non-significant in the silent fatty liver population. Future research is required to validate our findings and investigate the underlying mechanisms of these observations.

## Data Availability

All relevant data are within the manuscript and its Supporting Information files.

## Acknowledgments

Thank you to all the patients who participated in the study.

## Supporting information

**S1 Table. Risk of early liver fibrosis in HS with different metabolic dysfunction indices after PSM (blood pressure group)**.

**S2 Data. Raw Data**. The raw data is available in the S1 Table. Raw Data.

**S3 File. Ethical Approval Documentation**. The copy or scan of original ethical approval documentation and the proof of exemption from informed consent and their English translation are available in S3 File.

